# Indicators of Co-occurrence of Mood Disorder with Chronic Medical Conditions: Evidence from an Administrative Claims Data Analysis

**DOI:** 10.1101/2023.09.26.23296173

**Authors:** Karishma Chhabria, Trudy Millard Krause, Randa Hamden, Mbemba Jabbi

## Abstract

**Objective:** Mood disorder (including major depression and bipolar disorder) prevalence is over 10% and accounts for a significant share of global disease burden. Mental and physical illness are related, however, the association between mood disorders and acute/chronic disease subclasses remains poorly understood.

**Methods:** This observational cross-sectional study used administrative claims data from 6,709,258 adult enrollees with a full-year enrollment in the 2018 OPTUM Clinformatics® database. Data of enrollees with/without diagnoses of a mood disorder co-occurring with chronic comorbid conditions (defined by the Elixhauser Comorbidity Index) using the International Classification of Diseases (ICD-10) were analyzed by accounting for age, race, and ethnicity.

**Results:** Overall, the sample was predominantly non-Hispanic Caucasians (64.56%), with 48.59% females and a mean age of 43.54 years±12.46 years. The prevalence of mood disorders was 3.71% (248,890), of which 0.61% (n=40,616) had bipolar disorders and 3.10% (n=208,274) had Major Depressive Disorder (MDD). Logistic regression odds ratios revealed a strong association between mood disorder diagnoses and peptic ulcers (2.11; CI=2.01-2.21), weight loss (2.53; CI, 2.46-2.61), renal failure (2.37, CI = 2.31-2.42), peripheral vascular disease (2.24; CI=2.19-2.30), and pulmonary circulation disorder (1.77; CI=1.70-1.84).

**Conclusions:** Overall, mood disorders were associated with vascular and cardiac chronic medical conditions, suggesting a possible pathophysiological link between these conditions. The results highlight the importance of understanding the prevalence of co-occurring mood and medical conditions and may inform novel biological diagnostics and future identification of mechanisms for multimorbidity.

## INTRODUCTION

Mood disorders have a lifetime prevalence of over 10% worldwide (Murray et al., 2012; Collins et al., 2011; Kessler et al., 2007; Kessler et al., 2008; Plana-Ripoll et al., 2019). Mood disorders present a substantial share of the global disease burden as measured with disability-adjusted life years (DALYs) (Saloni Dattani, 2021; Whiteford et al., 2016). Globally, mood disorders affect approximately 310 million people, of which 264 million were documented to have a major depressive disorder (MDD) and 46 million had bipolar disorders (Dattani et al. 2023). The negative impact of mood disorders and co-occurring mental disorders on global disease burden and DALYs, a metric that considers both disorder-associated mortality and years lived with disability. Mood disorders also accounts for many premature mortalities, including close to a million suicides annually across the world (Evans et al., 2005; Ferrari et al., 2013; Momen et al., 2020; Weye et al., 2020). The hallmark of mood disorders (including major depressive disorder (MDD) and bipolar disorders) is debilitating depressive episodes that can co-occur with medical diseases like cardiovascular disease, autoimmune disease, and metabolic diseases such as diabetes (Momen et al., 2020). Although evidence suggests a strong association between mood disorders and co-occurring acute/chronic medical disease diagnoses (Evans et al., 2005; Ormel et al., 2007; Weye et al., 2020), the specific nature of the associations between mood disorders and subtypes of acute/chronic conditions remains understudied.

Indeed, several studies have separately assessed the association between MDD or bipolar disorder and other acute/chronic medical conditions (see comprehensive review by Evans et al.) (Evans et al., 2005; Machado et al., 2018). In a systematic review and meta-analysis of 40 articles, it was found that the risk for a diagnosis of depression was twice more likely for people with multimorbidity, the presence of two or more chronic conditions, in comparison to people without multimorbidity (Read et al. 2017). Furthermore, this observed risk for a depression diagnosis was three times greater for people with multimorbidity compared to those without any chronic physical condition (Read et al. 2017). However, evidence of an integrative cross-association between MDD and bipolar disorder with acute/chronic medical conditions is sparse.

Given that the diagnoses of MDD and bipolar disorder are often interrelated such that individuals initially diagnosed with MDD can subsequently have their diagnosis changed/rectified to be either bipolar disorder Type II or bipolar disorder type I, examining the association between mood disorders (which includes MDD and bipolar) and other conditions is therefore essential for the comprehensive understanding of the biological complexities underlying comorbid conditions. Depressive symptoms account for a significant disease burden for both MDD and bipolar disorders, and there is a clear need for a broader mood disorder-focused approach to understanding the pathological complexities of these prevalent conditions at the population level. Here, we studied the association of mood disorders with acute/chronic medical condition subtypes in a population of almost 7 million U.S. private insurance policyholders. Our study used administrative claims data to investigate mood disorders’ (including separate analysis for MDD and bipolar disorder) differential association and co-existence with acute/chronic disease comorbidities as defined by the Elixhauser’s comorbidity index (Elixhauser et al., 1998).

## METHODS

### Study Design and Population

We performed a retrospective observational study using OPTUM Clinformatics® Data Mart (CDM) to capture one year of claims data in 2018 (from January 1^st^ to December 31^st^). Our outcome of interest was defined as those with or without MDD, those with or without bipolar disorder, and a combination of MDD and bipolar disorder (referred to here as mood disorder). We compared our outcomes to those with and without chronic medical comorbidities as defined by Elixhauser’s comorbidity index (Elixhauser et al., 1998). The study sample included 6,709,258 adult enrollees between the ages of 21-64 years who were continuously enrolled for all 12 months of 2018 in OPTUM CDM.

OPTUM Clinformatics® Data Mart (CDM) is a statistically de-identified, HIPAA-complaint, closed system of insurance administrative claims, including patient enrollment; physician, facility, and pharmacy claims; and lab results from the largest health insurer in the U.S. Administrative claims data are collected for billing purposes hence data reported here are indicative of healthcare encounters with a recorded International Classification of Disease 10^th^ Edition (ICD-10) diagnosis codes. This database was accessed through The University of Texas Health Sciences Center Houston (UTHSC-H) School of Public Health Center for Health Care Data (CHCD). The UTHSC-H institutional review board approved the study. Since this study utilized de-identified administrative claims data via secondary data analysis, informed consent was not obtained. The data is available directly through OPTUM Clinformatics® Data Mart

### Measurements

A mood disorder diagnosis claim was assessed using ICD-10 diagnostic codes that capture the presence of bipolar disorder (includes bipolar I and II disorder), or MDD, during either inpatient or outpatient care encounters in 2018 (Appendix 2). Insurance claims data are collected for billing purposes; hence, claims evaluated in this study were recorded healthcare encounters such as diagnostic records from a psychiatrist or psychologist. We evaluated the frequency of healthcare encounters for MDD and bipolar disorder in the claims. Enrollees included in the bipolar disorder group were those with a definitive bipolar I or bipolar II disorder diagnosis (obtained from the claims data through healthcare encounters) in the study period. Enrollees that were categorized as having MDD were those who had a definitive diagnosis of MDD, regardless of a co-existing bipolar disorder diagnosis, with the exception that the frequency of MDD-related healthcare encounters would be greater than bipolar disorder. There were 2,594 enrollees with an equal frequency of claims (i.e., an equal number of healthcare encounters) for a diagnosis of MDD and bipolar disorder. Because these enrollees could not be definitively assigned to either group, they were excluded from our analysis.

As defined by the Elixhauser Comorbidity Index, acute/chronic comorbidities include 31 indicators that reflect acute/chronic medical conditions with significant short or long-term consequences on physical health (Elixhauser et al., 1998; Kim et al., 2018; Menendez et al., 2015; van Walraven et al., 2009). We assessed 24 indicators: blood loss anemia, cardiac arrhythmia, chronic pulmonary disease, coagulopathy, congestive heart failure, deficiency anemia, diabetes complicated, diabetes uncomplicated, fluid conditions, HIV/AIDS, hypertension, hypothyroidism, liver disease, lymphoma, metastatic cancer, obesity, peptic ulcers, peripheral vascular disorder, pulmonary circulation disorder, rheumatoid arthritis, renal failure, tumor without metastasis, valvular disease, and weight loss. We combined hypertension complicated and uncomplicated into a single indicator called hypertension. To account for additional factors that could influence the risk of study outcomes, we assessed demographics such as age, sex, and race/ethnicity as recorded in the patient’s enrollment file and included these variables in the analysis.

Adults with neurological or another primary diagnosis of a psychiatric condition, namely paralysis, neurological conditions, drug/alcohol use disorder diagnosis, and paralysis were excluded. Adults institutionalized in long-term care facilities were also excluded from the study due to the neurobiological cross-link between these indicators and study outcomes. (Appendix 1) (Devanand et al., 2022) (Collins et al., 2011)..

### Statistical Analysis

We performed descriptive statistics by first evaluating means, standard deviations, and prevalence of each co-existing acute/chronic comorbid medical condition across mood disorder subtypes. Unadjusted and adjusted odds ratios (OR) and confidence intervals (CI) were then calculated using binary variables for the presence or absence of exposure (acute and chronic medical conditions) and outcome diagnosis (pooled mood disorders, bipolar disorders, and MDD separately). Multivariable logistic regression models were used to evaluate the association of 1) bipolar disorder, 2) MDD, and 3) mood disorders based on comorbid conditions while accounting for age, sex, and race/ethnicity. Comorbid medical conditions that showed the top 5 highest prevalence ratios, i.e., a ratio calculated using prevalence rates from the overall population and mood disorder subtype, were included in the logistic models.

To assess the effects of age, we categorized age into four groups: 21-30 years, 31-40 years, 41-50 years, and 41-64 years for descriptive statistics but used it as a continuous variable in the logistic regression. We also included an interaction term between sex and age in our logistic models. Three separate models were run for each outcome: bipolar disorder, MDD, and mood disorders. The goodness of fit statistics and model convergence status were evaluated to assess model significance. Statistical significance was set at 0.05 apriori, and statistical analysis was performed using SAS, version 9.4 (SAS Institute Inc).

## RESULTS

From the 2018 Optum enrollment records, 6,709,258 enrollees met our inclusion criteria. The final sample of included enrollees were majority non-Hispanic Caucasians (64.56%), with a mean age of 43.54 (SD = 12.46) years. Table 1 presents the demographics and ethnicity of the included sample, of which 3.71% had a mood disorder diagnoses (n = 248,890), with the mood disorder sample comprising 0.61% bipolar disorder (n = 40,616) and 3.10% MDD (n = 208,274) of the total sample. Table 2 presents descriptive statistics of each chronic condition by study outcomes. We found that most of the included individuals in the non-mood disorder sample had no comorbid conditions (63.80%), while 36.20% had one or more of the identified comorbid medical conditions. Further, 64.56% of patients with a mood disorder had at least one comorbid medical condition. The most prevalent medical condition was hypertension, which affected 1,214,122 people (i.e., 18.97% of the non-mood disorder sample) (Table 2). Hypertension was also present in 34.46% of the mood disorder sample, of which 35.02% also had bipolar disorder, while 34.35% of the MDD sample had hypertension (Table 2).

**Table 1:** Demographic characteristics.

|  | **Non-mood disorder** | **Bipolar disorder** | **MDD** | **Mood disorder** |
| --- | --- | --- | --- | --- |
| N (%) | 6,460,378 | 40,616 (0.61) | 208,274 (3.10) | 248,890 (3.71) |
| Age (mean, SD) | 43.54 (12.46) | 45.54 (12.42) | 45.73 (12.64) | 45.70 (12.61) |
| Age |  |  |  |  |
| 21-30 years | 1,254,515 (19.42) | 6,384 (15.72) | 33,175 (15.93) | 39,559 (15.89) |
| 31-40 years | 1,494,741 (23.14) | 7,747 (19.07) | 39,927 (19.17) | 47,574 (19.15) |
| 41-50 years | 1,494,846 (23.14) | 9,615 (23.67) | 46,528 (22.34) | 56,143 (22.56) |
| 51-64 years | 2,216,266 (34.31) | 16,870 (41.54) | 88,644 (42.56) | 105,514 (42.39) |
| Sex |  |  |  |  |
| Male | 3,371,768 (52.19) | 13,275 (32.68) | 64,156 (30.80) | 77,431 (31.11) |
| Female | 3,088,600 (47.81) | 27,341 (67.32) | 144,118 (69.20) | 171,495 (68.89) |
| Race/ethnicity |  |  |  |  |
| Asian | 399,277 (6.18) | 763 (1.88) | 44,10 (2.12) | 5,173 (2.08) |
| African ancestry | 640,576 (9.92) | 4,084 (10.06) | 17,656 (8.48) | 21,740 (8.73) |
| Hispanic | 901,773 (13.96) | 3,535 (8.70) | 20,556 (9.87) | 24,091 (9.68) |
| Caucasian | 4,145,930 (64.17) | 30,193 (74.34) | 155,327 (7.45) | 185,520 (74.54) |
| Missing | 372,821 (5.77) | 2,041 (5.03) | 103,25 (4.95) | 12,364 (4.97) |

MDD: Major Depressive Disorder
SD: Standard Deviation

**Table 2:**
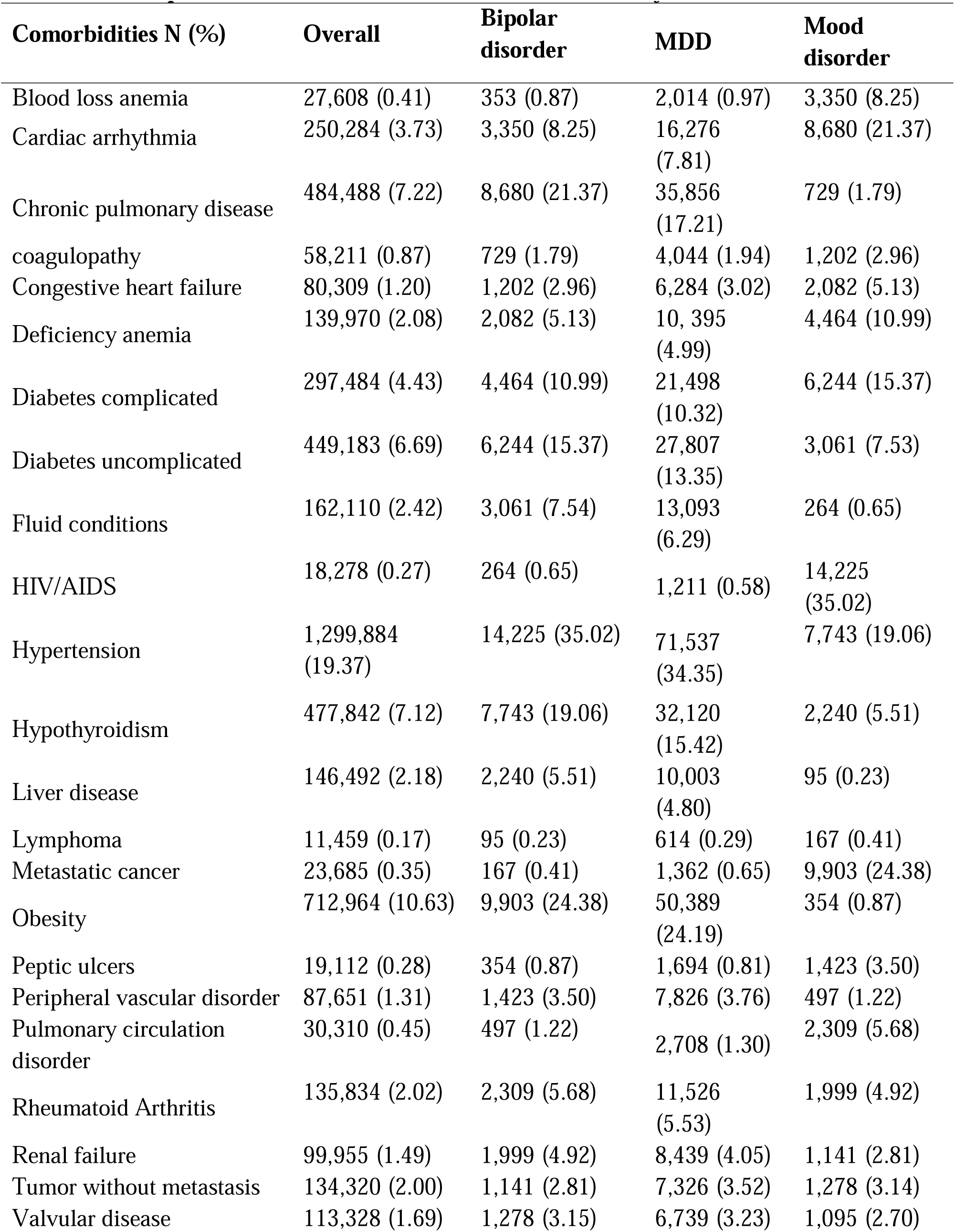
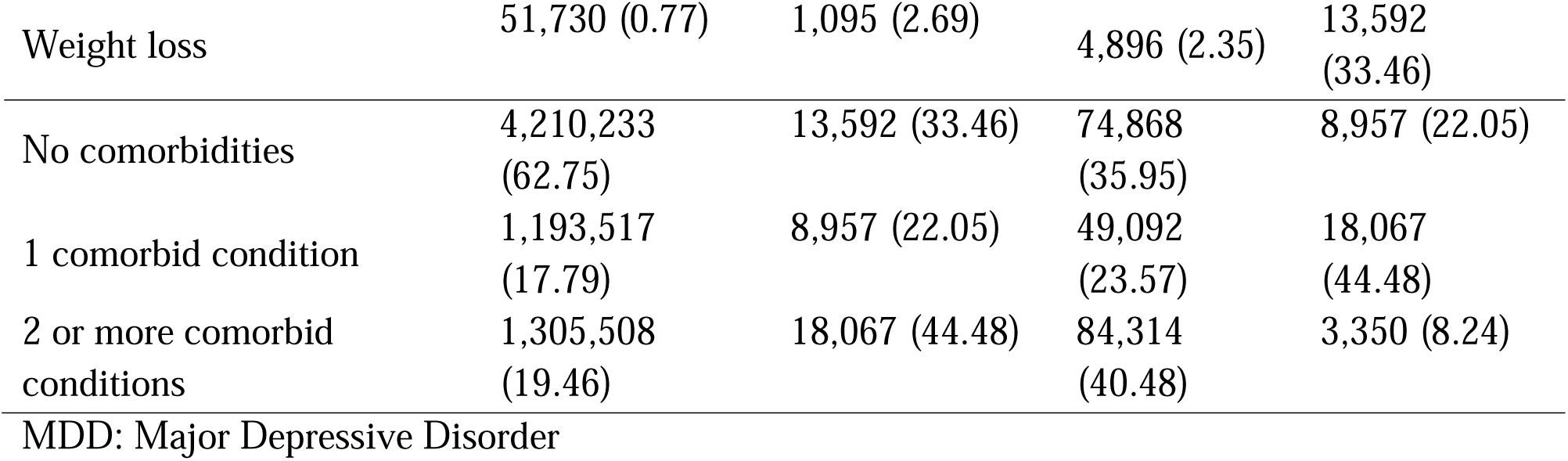
Descriptive statistics of chronic comorbid conditions by outcome variable.

| **Comorbidities N (%)** | **Overall** | **Bipolar disorder** | **MDD** | **Mood disorder** |
| --- | --- | --- | --- | --- |
| Blood loss anemia | 27,608 (0.41) | 353 (0.87) | 2,014 (0.97) | 3,350 (8.25) |
| Cardiac arrhythmia | 250,284 (3.73) | 3,350 (8.25) | 16,276 (7.81) | 8,680 (21.37) |
| Chronic pulmonary disease | 484,488 (7.22) | 8,680 (21.37) | 35,856 (17.21) | 729 (1.79) |
| coagulopathy | 58,211 (0.87) | 729 (1.79) | 4,044 (1.94) | 1,202 (2.96) |
| Congestive heart failure | 80,309 (1.20) | 1,202 (2.96) | 6,284 (3.02) | 2,082 (5.13) |
| Deficiency anemia | 139,970 (2.08) | 2,082 (5.13) | 10,395 (4.99) | 4,464 (10.99) |
| Diabetes complicated | 297,484 (4.43) | 4,464 (10.99) | 21,498 (10.32) | 6,244 (15.37) |
| Diabetes uncomplicated | 449,183 (6.69) | 6,244 (15.37) | 27,807 (13.35) | 3,061 (7.53) |
| Fluid conditions | 162,110 (2.42) | 3,061 (7.54) | 13,093 (6.29) | 264 (0.65) |
| HIV/AIDS | 18,278 (0.27) | 264 (0.65) | 1,211 (0.58) | 14,225 (35.02) |
| Hypertension | 1,299,884 (19.37) | 14,225 (35.02) | 71,537 (34.35) | 7,743 (19.06) |
| Hypothyroidism | 477,842 (7.12) | 7,743 (19.06) | 32,120 (15.42) | 2,240 (5.51) |
| Liver disease | 146,492 (2.18) | 2,240 (5.51) | 10,003 (4.80) | 95 (0.23) |
| Lymphoma | 11,459 (0.17) | 95 (0.23) | 614 (0.29) | 167 (0.41) |
| Metastatic cancer | 23,685 (0.35) | 167 (0.41) | 1,362 (0.65) | 9,903 (24.38) |
| Obesity | 712,964 (10.63) | 9,903 (24.38) | 50,389 (24.19) | 354 (0.87) |
| Peptic ulcers | 19,112 (0.28) | 354 (0.87) | 1,694 (0.81) | 1,423 (3.50) |
| Peripheral vascular disorder | 87,651 (1.31) | 1,423 (3.50) | 7,826 (3.76) | 497 (1.22) |
| Pulmonary circulation disorder | 30,310 (0.45) | 497 (1.22) | 2,708 (1.30) | 2,309 (5.68) |
| Rheumatoid Arthritis | 135,834 (2.02) | 2,309 (5.68) | 11,526 (5.53) | 1,999 (4.92) |
| Renal failure | 99,955 (1.49) | 1,999 (4.92) | 8,439 (4.05) | 1,141 (2.81) |
| Tumor without metastasis | 134,320 (2.00) | 1,141 (2.81) | 7,326 (3.52) | 1,278 (3.14) |
| Valvular disease | 113,328 (1.69) | 1,278 (3.15) | 6,739 (3.23) | 1,095 (2.70) |
| Weight loss | 51,730 (0.77) | 1,095 (2.69) | 4,896 (2.35) | 13,592<br>(33.46) |
| No comorbidities | 4,210,233<br>(62.75) | 13,592 (33.46) | 74,868<br>(35.95) | 8,957 (22.05) |
| 1 comorbid condition | 1,193,517<br>(17.79) | 8,957 (22.05) | 49,092<br>(23.57) | 18,067<br>(44.48) |
| 2 or more comorbid<br>conditions | 1,305,508<br>(19.46) | 18,067 (44.48) | 84,314<br>(40.48) | 3,350 (8.24) |
| MDD: Major Depressive Disorder |  |  |  |  |

### Prevalence Ratio Analyses

Using the prevalence ratio calculation, we found the top 5 most prevalent comorbid medical conditions for bipolar disorders included weight loss (3.49), renal failure (3.30), fluid conditions (3.11), and peptic ulcers (3.05). Similar prevalence ratio calculations revealed that the top 5 most prevalent comorbid medical conditions for MDD included weight loss (3.04), chronic pulmonary circulation disorder (2.87), peripheral vascular disorder (2.85), and rheumatoid arthritis (2.73).

Based on prevalence ratio calculation, the top 5 prevalent comorbid medical conditions we found to be associated with mood disorders included weight loss (3.12), peptic ulcers (2.88), pulmonary circulation disorder (2.85), peripheral vascular disorder (2.84) and renal failure (2.81).

### Multivariable Logistic Regression Analyses

Multivariate logistic regression met the criteria for convergence and goodness of fit for each of the models: bipolar disorders (percent concordant = 56.00), MDD (percent concordant = 63.70), and pooled mood disorders group (percent concordant = 63.80).

As shown in Table 3, the adjusted logistic regression model revealed a significant association between bipolar disorder and the top 5 conditions, including peptic ulcers (OR, 1.61; 95% CI, 1.45-1.81), weight loss (OR, 2.02, 95% CI, 1.89-2.15), renal failure (OR, 2.10, 95%CI = 1.99 – 2.20), fluid conditions (OR, 1.87; 95% CI, 1.79 – 1.95), and chronic pulmonary disease (OR, 2.70; 95% CI, 2.63-2.77). The model further revealed that being Caucasian, of advanced age (OR=1.00, 95% CI = 1.00 – 1.00), and being female (OR = 2.05, 95% CI = 2.01-2.10) were associated with bipolar disorders diagnosis. The interaction term between age and sex is also significantly associated with bipolar disorders diagnosis (OR=2.05, 95% CI = 2.01 – 2.10, p-value = 0.001) (Table 3).

**Table 3:**
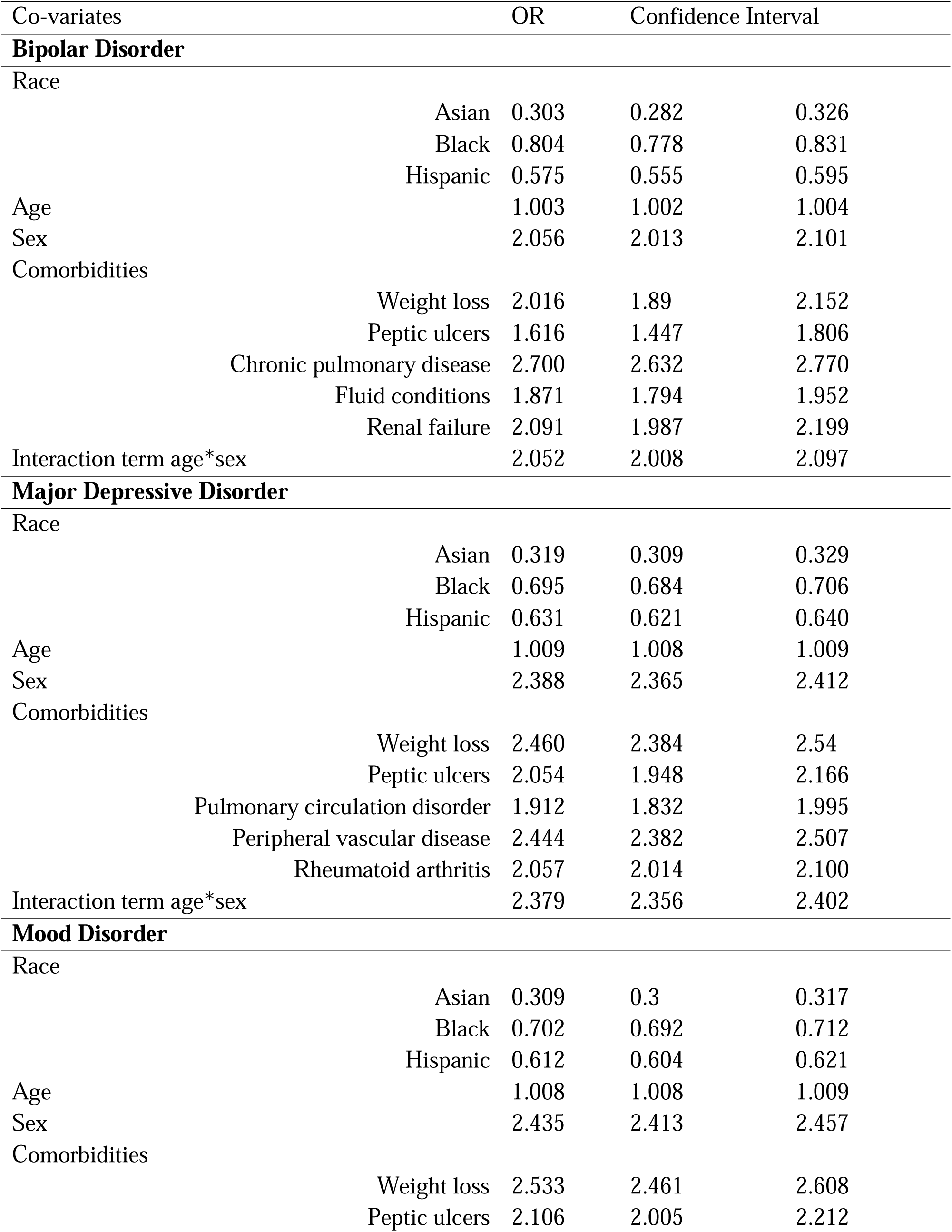
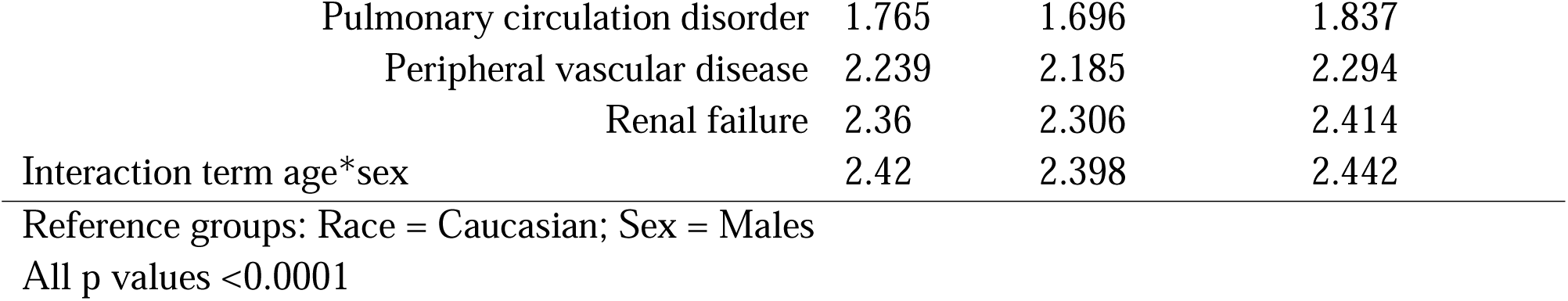
Adjusted Odds Ratio.

| Co-variates |  | OR | Confidence Interval |  |
| --- | --- | --- | --- | --- |
| Bipolar Disorder |  |  |  |  |
| Race |  |  |  |  |
|  | Asian | 0.303 | 0.282 | 0.326 |
|  | Black | 0.804 | 0.778 | 0.831 |
|  | Hispanic | 0.575 | 0.555 | 0.595 |
| Age |  | 1.003 | 1.002 | 1.004 |
| Sex |  | 2.056 | 2.013 | 2.101 |
| Comorbidities |  |  |  |  |
|  | Weight loss | 2.016 | 1.89 | 2.152 |
|  | Peptic ulcers | 1.616 | 1.447 | 1.806 |
|  | Chronic pulmonary disease | 2.700 | 2.632 | 2.770 |
|  | Fluid conditions | 1.871 | 1.794 | 1.952 |
|  | Renal failure | 2.091 | 1.987 | 2.199 |
| Interaction term age*sex |  | 2.052 | 2.008 | 2.097 |
| Major Depressive Disorder |  |  |  |  |
| Race |  |  |  |  |
|  | Asian | 0.319 | 0.309 | 0.329 |
|  | Black | 0.695 | 0.684 | 0.706 |
|  | Hispanic | 0.631 | 0.621 | 0.640 |
| Age |  | 1.009 | 1.008 | 1.009 |
| Sex |  | 2.388 | 2.365 | 2.412 |
| Comorbidities |  |  |  |  |
|  | Weight loss | 2.460 | 2.384 | 2.54 |
|  | Peptic ulcers | 2.054 | 1.948 | 2.166 |
|  | Pulmonary circulation disorder | 1.912 | 1.832 | 1.995 |
|  | Peripheral vascular disease | 2.444 | 2.382 | 2.507 |
|  | Rheumatoid arthritis | 2.057 | 2.014 | 2.100 |
| Interaction term age*sex |  | 2.379 | 2.356 | 2.402 |
| Mood Disorder |  |  |  |  |
| Race |  |  |  |  |
|  | Asian | 0.309 | 0.3 | 0.317 |
|  | Black | 0.702 | 0.692 | 0.712 |
|  | Hispanic | 0.612 | 0.604 | 0.621 |
| Age |  | 1.008 | 1.008 | 1.009 |
| Sex |  | 2.435 | 2.413 | 2.457 |
| Comorbidities |  |  |  |  |
|  | Weight loss | 2.533 | 2.461 | 2.608 |
|  | Peptic ulcers | 2.106 | 2.005 | 2.212 |

|  |  |  |  |
| --- | --- | --- | --- |
| Pulmonary circulation disorder | 1.765 | 1.696 | 1.837 |
| Peripheral vascular disease | 2.239 | 2.185 | 2.294 |
| Renal failure | 2.36 | 2.306 | 2.414 |
| Interaction term age*sex | 2.42 | 2.398 | 2.442 |

Reference groups: Race = Caucasian; Sex = Males
All p values <0.0001

As shown in Table 3, the adjusted logistic model revealed an association of MDD with the top 5 prevalence ratio conditions, including peptic ulcers (OR, 2.05; 95% CI, 1.95-2.17), weight loss (OR, 2.46; 95% CI, 2.38-2.54), peripheral vascular disease (OR, 2.44; 95% CI, 2.38-2.51), pulmonary circulation disorder (OR, 1.91; 95% CI, 1.83-2.00), and rheumatoid arthritis (OR, 2.06; 95% CI, 2.01-2.10). Further, the adjusted logistic regression model showed that being Caucasian, older age, and female increased the likelihood of having MDD, while sex interacted with age such that older female adults had a higher likelihood of having MDD (OR, 2.38; 95% CI, 2.36-2.40).

The adjusted logistic regression model shows the top 5 prevalence ratio comorbid medical conditions for mood disorders to include peptic ulcers (OR=2.11, 95% CI = 2.01 – 2.21), weight loss (OR=2.53, 95% CI = 2.46 – 2.61), renal failure (OR=2.37, 95% CI = 2.31 – 2.42), pulmonary circulation disorder (OR=1.77, 95% CI = 1.70 – 1.84) and peripheral vascular disease (OR=2.24, 95% CI = 2.19 – 2.30) (Table 5). Furthermore, Caucasians were found to have the highest likelihood of a mood disorder diagnosis compared to other races (p < 0.001). Females were 2.4 times more likely to have a mood disorder diagnosis (OR=2.43, 95% CI = 2.41 – 2.46), and the observed sex effect interacted with age in that older females were more likely to have a mood disorder diagnosis (OR=2.42, 95% CI = 2.40 – 2.44).

## DISCUSSION

Mood disorders affect over ten percent of the population over the lifespan and frequently co-occur with comorbid medical conditions (Dalby et al. 2022). In this observational study, we used insurance claims data to understand the associations (or co-occurrence) between mood disorders and chronic comorbid conditions using records of healthcare encounters via healthcare insurance claims. Although our results are strictly observational and therefore can not address causality based on the analysis and overall approach of our study, our results did reveal strong associations between mood disorders with 1) peptic ulcers, 2) weight loss, 3) renal failure, 4) pulmonary circulation disorder, and 5) peripheral vascular disorder. These vascular and cardiac disease associations with mood disorder diagnoses align with earlier work (Evans et al., 2005; Momen et al., 2020; Momen et al., 2022). Bipolar disorder diagnosis was associated with 1) peptic ulcers, 2) weight loss, 3) renal failure, 4) fluid conditions, and 5) chronic pulmonary disease. MDD, on the other hand, was associated with 1) peptic ulcers, 2) weight loss, 3) peripheral vascular disease, 4) pulmonary circulation disorder, and 5) rheumatoid arthritis (Momen et al., 2020; Momen et al., 2022). These findings align with previous results showing strong associations between mood disorders and ulcers, weight loss, cardiovascular diseases (Evans et al., 2005; Momen et al., 2022), and pulmonary diseases (Panagioti et al. 2014). For instance, Panagioti et al. showed that over one-third of individuals with chronic obstructive pulmonary disease suffer comorbid mood and anxiety disorders (Panagioti et al. 2014). In another independent report, the number of comorbidities between chronic diseases and mental disorders has been shown to influence overall treatment outcomes (Garvey & Criner 2018).

In our large-scale study sample, we found a fivefold higher prevalence of MDD in the overall sample than in bipolar disorder. Within the pooled mood disorders sample, the association between weight loss was the highest observed somatic disease comorbidity, in line with weight instability being a core somatic symptom of mood disorders. Although MDD is more prevalent than bipolar disorder, bipolar disorder is documented to be more heritable (i.e., bipolar disorder has a higher familial diagnostic occurrence than MDD) (McIntyre et al., 2020; Mendlewicz, 2009; Otte et al., 2016). Our findings of convergent associations between comorbid medical conditions and both bipolar disorders and MDD (i.e., mood disorders) suggest possible comorbidity overlaps across mood disorders and sub-classes of acute/chronic diseases when these medical conditions are classified according to biological/organ-specific dysfunctions (see Tylee D.S. et al. 2022 as an example) (Tylee et al., 2022).

There likely are many possible causal or biological mechanisms driving the observed comorbidities. For instance, environmental pathogens, such as infective agents and socially adverse as well as naturally adverse experiences, can cumulatively cause excessive inflammatory responses that can lead to or exacerbate various chronic and mental diseases (Baltrusch et al. 1991; von Kanel 2012; Stuller et al. 2012; Nikiphorou et al. 2019; Jackson and Jabbi, 2022; Brewerton 2022). Of note, four of the top 5 most likely comorbid medical conditions were the same for bipolar disorder and MDD. Such overlapping associations may echo shared mood disorders etiology in that psychological and environmentally adverse and stressful experiences can trigger emergent mood disorders and cardiovascular and other somatic conditions (Jabbi and Nemeroff, 2019; Baltrusch et al. 1991; von Kanel, 2012; Stuller et al. 2012; Nikiphorou et al. 2019; Jackson and Jabbi, 2022; Brewerton, 2022). In addition, this idea aligns with a recent study showing that a history of mental stress, with and without conventional stress, was associated with a significantly increased risk for myocardial ischemia with cardiovascular events (Vaccarino et al., 2021).

Together, our findings of comorbid circulatory and cardiovascular disease with mood disorders, in addition to peptic ulcers, weight loss, and renal failure, may have implications for a better understanding of disease outcomes in light of evidence showing increased mortality in depression when co-occurring with cardiovascular and other diseases (Frasure-Smith et al., 1995).

For instance, one hypothesis states that a bidirectional link exists between mood disorders and acute/chronic comorbid conditions (Evans et al., 2005). An alternative hypothesis suggests that mood disorders can affect the course of medical diseases like ischemic heart disease (Ferrari et al., 2013; Frasure-Smith et al., 1995; Headrick et al., 2017) by increasing overall disease burden and negative prognostic and severe disease outcome over time (Frasure-Smith et al., 1995; Manning, 2005; McGuire et al., 2002), including increasing the risk for mortality (Insel and Charney, 2003; Struble et al., 2014) and suicide (Sanna et al., 2014). In line with these models, we found a robust association between mood disorders and gastrointestinal (peptic ulcers), metabolic (weight loss), and acute and chronic circulatory/renal and cardiovascular diseases in a large adult sample of over 6.7 million individuals. These findings indicate a possible additive contribution of mood disorders and acute/chronic medical comorbidities. However, how the observed comorbidities emerge, what their combined disease burden is (Ferrari et al., 2013), and what cumulative impact these measures can have on overall morbidity and mortality indicators, including suicide rates over time, needs to be addressed with future longitudinal studies (Ferrari et al., 2013; McIntyre et al., 2020).

### Limitations

This study needs to be interpreted in light of its limitations. *First*, our study examined a one-year window of private insurance claims from January 2018 to December 2018. This focus on 1-year data limits the generalizability of our findings over longer durations. Using a private insurance dataset also limits the generalizability of our results across populations without private insurance, such as Medicaid and the uninsured.

Furthermore, focusing on active diagnoses without accounting for symptom history, including mood episodic history and remission status, limits the study’s ability to shed light on the longer-term implications of the observed comorbidities. However, the use of administrative claims data resulting from healthcare encounters highlights the definitive diagnosis of these conditions as opposed to patient-reported outcomes, which may be subject to bias. Here, we focused only on adults aged 21-64, excluding children, adolescents, post-adolescents, and individuals 65 and older. This selective age window limits the interpretation of our findings beyond the studied age range. *Second*, disease comorbidities can have complex and varying etiological and biological relationships. This complexity implies that more data on the possible biological and etiological correlates of such comorbidities, coupled with data on the time-relevant relationships between the comorbid disorders in terms of what diagnostic subclass emerges first, are needed to better understand the mechanistic aspects of our observed comorbidities. Moreover, our work strictly addresses associations and not causations. However, future studies of the causal relationships between our observed associations using time series of diagnostic data would shed more light on the directional influences of our observed associations. *Third*, we excluded all mental illness and neurological/neuropsychiatric comorbidities given the proximate cross-link between these conditions and mood disorders with brain systems and endpoint behavioral symptoms. While this is necessary, it limits the possibility of assessing if comorbidities between mood disorders and other neuropsychiatric conditions could moderate further prevalence of comorbid mood disorders with mental and medical conditions. Fourth, an individual could be diagnosed with MDD and later develop bipolar disorder. As is commonly known, a one-time diagnostic healthcare record for MDD and a follow-up diagnosis of bipolar disorder after the first onset of mania cannot be identified and excluded in this cohort due to the nature of the claims. However, only the most recent mood disorder diagnosis within the studied period was included. We have additionally integrated both MDD and bipolar disorder into a mood disorder diagnostic group, an approach we adopted to minimize this limitation given the close diagnostic relationship between the two conditions. *Finally*, we think that leading models positing that mood disorders and medical conditions are bi-directionally associated or that mood disorders can negatively impact medical disease prognoses and outcomes are more complementary than mutually exclusive. Therefore, our study captures associations between the diagnostic presence of comorbid medical conditions and mood disorders in a large sample of over 6.7 million U.S. adults. It cannot assess whether the mood disorder preceded the medical disease onsets and whether the disease course of the medical conditions was affected by the presence of mood disorders. By including 6.7 million individuals in our study and identifying bodily systems-related comorbidities with mood disorder in one year capture using the Elixhauser Comorbidity Index, our observations of mood disorder-medical comorbidities provide further biological insights into the possible pathophysiological underpinnings of complex comorbidities.

## Conclusion

This diagnostic association study used the Optum private insurance claims data to assess the association between mood disorders and medical conditions organized in comorbid medical conditions of organ systems. We included over 6.7 million insured individuals using the Elixhauser Comorbidity Index. Although the Elixhauser comorbidity index uses a dichotomous categorization of whether a certain comorbidity is present or absent, the index has been validated to have predictive values for healthcare consumption, hospitalization, and even mortality risk (Elixhauser et al., 1998). Therefore, our study identifies a mood disorder association with comorbid medical conditions such as peptic ulcers, weight loss, and cardiovascular/circulatory diseases. The observed mood disorders’ associations with acute and chronic medical conditions are compelling. They could contribute to improving diagnostic practices currently designed to fractionate mood/mental disorders from co-occurring conditions and provide a framework for predicting morbidity and mortality outcomes. Our results add to the current evidence highlighting the need to reduce the stigma associated with mood disorders and encourage a paradigm shift towards considering the possible clinical and physiological heterogeneity of mood disorders and other medical conditions at various mechanistic and etiological levels. In sum, using a large population-based investigation, our results highlight the importance of understanding prevalent comorbidities between mood disorders and other acute and chronic medical conditions.

## Data Availability

Data will be made available upon request to the UTHEalth Houston Data Center.

---

**Appendix 1:** ICD-10 codes for exclusion criteria.

| Code | Description |
| --- | --- |
| G10.x | Huntington's chorea · Huntington's dementia. |
| G11.x | Hereditary ataxia. |
| G12.x | Spinal Muscular Atrophy and Related Syndromes |
| G13.x | Systemic atrophies primarily affecting the central nervous system in diseases classified elsewhere. |
| G20.x | Parkinson's disease. |
| G21.x | Secondary parkinsonism. |
| G22.x | Secondary parkinsonism, unspecified |
| G35.x | multiple sclerosis |
| G36.x | Other acute disseminated demyelination |
| G37.x | Other demyelinating diseases of the central nervous system. |
| G25.4 | Drug-induced chorea |
| G25.5 | Other chorea |
| G31.2 | Degeneration of nervous system due to alcohol |
| G31.8 | Other specified degenerative diseases of the nervous system |
| G31.9 | Degenerative disease of the nervous system, unspecified |
| G93.1 | Anoxic brain damage, not elsewhere classified. |
| G39.4 | Other and unspecified encephalopathy. |
| R47.0 | Dysphasia and aphasia |
| G32.x | Other degenerative disorders of the nervous system in diseases are classified elsewhere. |
| G40.x | Epilepsy and recurrent seizures |
| G41.x | Status epilepticus |
| R56.x | Convulsions, not elsewhere classified |
| G04.1 | Tropical spastic paraplegia |
| G80.1 | Spastic diplegic cerebral palsy |
| G80.2 | Spastic hemiplegic cerebral palsy |
| G81.x | Hemiplegia and hemiparesis following unspecified cerebrovascular disease. |
| G82.x | Paraplegia (paraparesis) and quadriplegia (quadriparesis) |
| G83.0 | Diplegia of upper limbs |
| G83.1 | Monoplegia of lower limb |
| G83.2 | Monoplegia of upper limb. |
| G83.4 | Cauda equina syndrome |
| G83.9 | Paralytic syndrome, unspecified |
| G32.x | Other degenerative disorders of nervous system in diseases classified elsewhere |
| G93.1 | Anoxic brain damage, not elsewhere classified |
| G93.4 | Other and unspecified encephalopathy |
| R47.0 | Dysphasia and aphasia |
| F10.x | Alcohol related disorders |
| E52.x | Niacin deficiency [pellagra] |
| G62.1 | Alcoholic polyneuropathy |
| I42.6 | Alcoholic cardiomyopathy |
| K29.2 | Alcoholic gastritis |
| K70.0 | Alcoholic fatty liver |
| K70.3 | Alcoholic cirrhosis of liver |
| K70.9 | Alcoholic liver disease, unspecified |
| T51.x | Toxic effect of alcohol |
| Z50.2 | Alcohol rehabilitation |
| Z71.4 | Alcohol abuse counseling and surveillance |
| Z72.1 | Alcohol use |
| F20.x | Schizophrenia. |
| F22.x | Delusional Disorders |
| F23.x | Brief psychotic disorder |
| F24.x | Shared psychotic disorder |
| F25.x | Schizoaffective disorder, bipolar disorder type |
| F28.x | Other psychotic disorder not due to a substance or known physiological condition |
| F29.x | Unspecified psychosis not due to a substance or known physiological condition |
| F30.2 | Mania with psychotic symptoms |
| F11.x | Opioid related disorders. |
| F12.x | Cannabis abuse, uncomplicated |
| F13.x | Sedative, hypnotic or anxiolytic abuse, uncomplicated |
| F14.x | use of cocaine |
| F15.x | use of other stimulants, including caffeine |
| F16.x | use of hallucinogens |
| F18.x | use of inhalants |
| F19.x | use of other psychoactive substances and multiple drug use |
| Z71.5 | Drug abuse counseling and surveillance. |
| Z72.2 | Drug use |

**Appendix 2:** ICD-10 codes identifying mood disorder.

| Code | Description |
| --- | --- |
| **Bipolar Disorder** |  |
| F3110 | Bipolar disorder, current episode manic without psychotic features, unspecified |
| F3111 | Bipolar disorder, current episode manic without psychotic features, mild. |
| F3112 | Bipolar disorder, current episode manic without psychotic features, moderate |
| F3113 | Bipolar disorder, current episode manic without psychotic features, severe |
| F312 | Bipolar affective disorder, current episode manic with psychotic symptoms |
| F3130 | Bipolar disorder, current episode depressed, mild or moderate severity, unspecified. |
| F3131 | Bipolar disorder, current episode depressed, mild. |
| F314 | Bipolar affective disorder, current episode severe depression without psychotic symptoms. |
| F315 | Bipolar disorder, current episode depressed, severe, with psychotic features |
| F3160 | Bipolar disorder, current episode mixed, unspecified |
| F3161 | Bipolar disorder, current episode mixed, mild. |
| F3162 | Bipolar disorder, current episode mixed, moderate |
| F3163 | Bipolar disorder, current episode mixed, severe, without psychotic features. |
| F3164 | Bipolar disorder, current episode mixed, severe, with psychotic features. |
| F3170 | Bipolar disorder, currently in remission, most recent episode unspecified. |
| F3171 | Bipolar disorder, in partial remission, most recent episode hypomanic. |
| F3172 | Bipolar disorder, in full remission, most recent episode hypomanic |
| F3173 | Bipolar disorder, in partial remission, most recent episode manic. |
| F3174 | Bipolar disorder, in full remission, most recent episode manic. |
| F3175 | Bipolar disorder, in partial remission, most recent episode depressed |
| F3176 | Bipolar disorder, in full remission, most recent episode depressed. |
| F3177 | Bipolar disorder, in partial remission, most recent episode mixed |
| F3178 | Bipolar disorder, in full remission, most recent episode mixed. |
| F3181 | Bipolar II disorder. |
| F3132 | Bipolar disorder, current episode depressed, moderate. |
| **Major Depressive Disorder** |  |
| F330 | Major depressive disorder, recurrent, mild |
| F331 | Major depressive disorder, recurrent, moderate. |
| F332 | Recurrent depressive disorder, current episode severe without psychotic symptoms. A |
| F333 | Recurrent depressive disorder, current episode severe with psychotic symptoms. |
| F312 | Bipolar affective disorder, current episode manic with psychotic symptoms. |
| F3340 | Major depressive disorder, recurrent, in remission, unspecified |
| F3341 | Major depressive disorder, recurrent, in partial remission. |
| F3342 | Major depressive disorder, recurrent, in full remission. |
| F338 | Other recurrent depressive disorders. |
| F339 | Major depressive disorder, recurrent, unspecified. |

